# Risk factors for SARS-Cov-2 infection at a United Kingdom electricity-generating company: a test-negative design case-control study

**DOI:** 10.1101/2023.08.25.23294609

**Authors:** Charlotte E Rutter, Martie Van Tongeren, Tony Fletcher, Sarah A Rhodes, Yiqun Chen, Ian Hall, Nick Warren, Neil Pearce

## Abstract

**Objectives:** Identify workplace risk factors for SARS-Cov-2 infection, using data collected by a United Kingdom electricity-generating company.

**Methods:** Using a test-negative design case-control study we estimated the odds ratios (OR) of infection by job category, site, test reason, sex, vaccination status, vulnerability, site outage, and site COVID-19 weekly risk rating, adjusting for age, test date and test type.

**Results:** From an original 80,077 COVID-19 tests, there were 70,646 included in the final analysis. Most exclusions were due to being visitor tests (5,030) or tests after an individual first tested positive (2,968).

Women were less likely to test positive than men (OR=0.71; 95% confidence interval=0.58-0.86). Test reason was strongly associated with positivity and although not a cause of infection itself, due to differing test regimes by area it was a strong confounder for other variables. Compared to routine tests, tests due to symptoms were highest risk (94.99; 78.29-115.24), followed by close contact (16.73; 13.80-20.29) and broader-defined work contact 2.66 (1.99-3.56). After adjustment, we found little difference in risk by job category, but some differences by site with three sites showing substantially lower risks, and one site showing higher risks in the final model.

**Conclusions:** In general, infection risk was not associated with job category. Vulnerable individuals were at slightly lower risk, tests during outages were higher risk, vaccination showed no evidence of an effect on testing positive, and site COVID-19 risk rating did not show an ordered trend in positivity rates.

**Key messages:** *What is already known on this topic:*

- In the United Kingdom, there is now a considerable body of evidence showing occupational differences in Covid-19 infection and severity, but with understandable focus on high-risk industries like healthcare.
- Less is known about differences in risk of COVID-19 infection in other industries that do not involve directly working with the general public, in particular, there is relatively little evidence on the risks of transmission in the electricity-generating industry.

*What this study adds:*

- At this company, infection risk was not associated with job category after adjusting for test reason; however women were less likely to test positive than men and the risk was higher when there was a power outage, requiring more staff to visit the site in person.

*How this study might affect research, practice or policy:*

- The site risk rating showed a consistent (but modest) dose-response with infection risk, indicating that such risk rating may be useful for identifying “high risk” sites.
- This analysis demonstrates the importance of adjusting for both date of and reason for test, when prevalence and testing protocols differ over time.

## Introduction

From the outset of the Covid-19 pandemic there was considerable debate as to the role of occupational exposures in the transmission of infections, and the subsequent morbidity and mortality. Most deaths occurred in those over age 65 years, i.e., in those above the usual working age. However, there was still a substantial number of infections, hospital admissions, and deaths in those of working age.[1]

Thus, one year into the pandemic, Burdorf et al [2] commented that “there is scattered evidence that an individual’s type of job may contribute to the risk of becoming infected and, hence, to the mortality pattern in society”. Three years into the pandemic, Michaels et al [3] drew stronger conclusions, arguing that “Covid-19 is an occupational disease that sickened and killed countless workers”, but noted that Covid-19 has rarely been treated or tracked as an occupational disease by public health agencies, particularly in non-health-care workplaces. They also noted that most white-collar workers could work from home, and that perhaps greater priority might have been placed on making workplaces safe if this had not been the case.

In the United Kingdom, there is now a considerable body of evidence showing occupational differences in Covid-19 infection [4-6], severity [7], vaccination availability and uptake [8,9], mortality [1, 10-12], and mitigation strategies [13,6].

However, these studies have mostly considered all occupations together in agnostic analyses, with considerable breadth, but little depth. More in-depth analyses have generally focused on known “high risk” industries such as health care, social care, and other “essential occupations” involving regular contact with the general public.[14,15]

There have been more in-depth investigations in a plastics manufacturing plant [16] and an automotive manufacturing site [17] but these have been largely descriptive and have involved only a small number of Covid-19 cases.

To our knowledge, there has not previously been a specific study of electricity-generation workers. This industry is of considerable interest, since working from home is not a possibility for almost all operational staff, and there is little or no contact with the general public during working hours. Therefore, it provides an opportunity to investigate factors affecting workplace transmission in this specific environment.

In July 2020, government scientific advisers and key funders identified where the United Kingdom (UK) must increase research to respond to near term strategic, policy and operational needs, and ultimately improve resilience against COVID-19 through 2021 and beyond. Six COVID-19 National Core Studies (NCS) have been established to meet these needs, including the NCS project on transmission of the SARS-CoV-2, led by the UK’s Health and Safety Executive (HSE). This project is known as “PROTECT”: The Partnership for Research into Occupational, Transport and Environmental Covid NCS, which brings together more than 70 researchers from 16 different institutions.

One of the six key themes of the “PROTECT” project is to collect data from outbreak investigations in a range of workplaces to understand SARS-CoV-2 transmission risk factors, potential causes for COVID-19 outbreaks and the effectiveness of a range of measures to control and prevent these outbreaks. In addition to specific outbreak investigations conducted as part of PROTECT, some companies have been identified which succeeded in assembling some detailed data on testing in their workforces including relevant data on outbreaks they have experienced. One of these is a large electricity-generating company. We here report the findings of a test-negative design case-control study conducted using the data collected by this company. The main aim of these analyses was to investigate contextual-level, workplace subgroups and individual-level risk factors for SARS-Cov-2 infections.

## Methods

The large electricity-generating company which is the subject of this report, tested staff frequently on site throughout the course of the pandemic. The testing strategy and method varied over time and by facility. During some time-periods, all staff were tested routinely, whereas in other time-periods, most workers were only tested because they had symptoms, or were identified as a contact of a positive case. These practices also varied across sites, so that at any given time, some sites may have been testing routinely, while others were only doing symptomatic and contact testing. Reason for test was collected and categorised into 4 groups: testing due to symptoms (using a lower threshold than government recommendations), testing for close contacts (using government defined criteria), testing for broader-defined work contacts (as per company protocols) and routine testing.

Tests with a missing or inconclusive result were excluded, along with those from visitors (single tests), and any person with missing job type. We also excluded tests that were missing one of the following a priori confounders: age, sex, site, test date or test type.

We included a maximum of one test per day for each person. Where there were multiple tests in a day with different outcomes or reasons (only a small number were identified), we prioritised positive results over negative (as false positives are less common than false negatives), and test reason in order of strength of reason (i.e., symptoms, close contact, broader-defined work contact, screening, and then missing). For each person, we used tests only up to and including their first positive result. Thus, the analyses presented here relate to the risks of a first infection, and subsequent infections were not considered.

We used a test-negative design, in which positive tests (cases) were compared with negative tests (controls) during each quarter (3-month period). This approach is intended to control for factors that affect the propensity to be tested at different time points (e.g. changing testing protocols and recognition of symptoms). It also has the advantage of being feasible, since we only had access to the test data, and not to data on individuals who were not tested. It has been widely used for assessing vaccine effectiveness, both for COVID-19[18] and for other infection[19]. More recently, it has been used for assessing risk factors for COVID-19 infection.[20,21]Site risk rating was assessed by the Outbreak Management Team consisting of the company doctors, occupational health advisors and site representatives for each power station, approximately once a week, based on the background prevalence of disease and the number of cases on site. The risk rating from 0 to 5 determined the COVID-19 mitigation requirements (e.g. cleaning, PPE, testing, social distancing). At the higher risk ratings more than half the permanent workforce of the power stations were working remotely, face coverings were mandatory at all times inside, and enhanced contact tracing and isolation used a broader definition of a contact than in the national regulations. If there was no available risk rating for a particular week then the rating for the closest previous date was chosen. For sites with no risk rating assigned then the average risk rating across all sites was used.

An outage is a statutory shutdown, when a power station is offline, and maintenance can be undertaken. It is often a time with an increased number of external visitors/contractors to the site (sometimes doubling the number of people on site). We have a binary flag determining whether a test was taken during an outage at the relevant site.

Vulnerability status was determined by increased risk of severe disease or death from COVID-19 due to a pre-existing health condition as determined by the literature and was based on an employee’s request for assessment. We have a binary flag for identified vulnerable staff members, who followed different protocols for their own protection (such as home working practices).

Vaccination information was captured for most workers on a voluntary basis. Where we had date of vaccine then we could determine vaccine status at the time of the test, defining vaccine immunity as beginning 10 days after the vaccination date. Partial vaccination was defined as having received one vaccine (10 days or more prior) and full vaccination as two or more. Individuals without vaccination information were assumed unvaccinated as negative information was not captured. The Janssen vaccine which requires only one dose was not widely used in the UK and no single doses of this type were recorded in this study.

There were different types of tests available at different sites at different times and for different reasons. We categorised the tests as PCR LAMP (polymerase chain reaction loop-mediated isothermal amplification), other PCR (polymerase chain reaction), and LFT (lateral flow test). For PCR LAMP, all positives and 10% of negatives were then confirmed by PCR.

We fitted logistic regression models comparing tests with positive outcomes to those with negative outcomes adjusting for the time-period. There may have been more than one test per person during a time-period but unless the tests were very close together there would not be much dependency. In addition to the a priori confounders of age, sex, date of test, and test type, we considered the other available information detailed above as potential confounders or other explanatory variables of interest. Some of these factors were related, e.g., routine testing was more common during site outages. All potential confounders were included in the final model, provided there were no problems of collinearity or non-convergence of the model. Ordinal variables were tested for linear trend using a likelihood ratio test and included as either categorical or continuous based on the result. The analysis used Stata version 17.

## Results

From an original file of 80,077 tests there were 70,878 included in the analysis (Table 1). Most exclusions were due to being visitor tests (5,030) or being tests for an individual after they first tested positive (2,968, of which 433 [14.6%] were also positive).

**Table 1:** Test numbers and exclusions.

| Exclusion reason (in order) | Excluded tests | Remaining tests |
| --- | --- | --- |
| **Total** |  | 80,077 |
| Missing/invalid test outcome | 200 | 79,877 |
| Missing test date | 0 | 79,877 |
| Missing job category | 267 | 79,610 |
| Visitor job category | 5,030 | 74,580 |
| Missing site | 0 | 74,580 |
| Missing sex | 0 | 74,580 |
| Missing age group | 1 | 74,579 |
| Missing test type | 601 | 74,579 |
| Multiple tests in single day<sup>a</sup> | 132 | 74,447 |
| Tests after first testing positive | 2,968 | 70,878 |

<sup>a</sup> duplicates deleted; 17 people had both negative and positive tests of which one positive test was kept. Of those with matching outcomes, 31 people had different test reasons of which the highest priority one was kept in this order: symptoms, close contact, broader-defined work contact, screening, missing reason.

Table 2 shows the demographic characteristics of the study participants. Almost 90% of the workers tested were men. There was a wide spread of ages from under 20 to over 70 and the median age group was 41-45. The largest proportion of workers were external contractors (53%), followed by engineering (16%) and operations (13%). Jobs were spread in a variety of locations, most at power stations, and there were many more tests per person at power stations 3, 7 and 8 (around 10 per person on average) than at other sites (between 2-5 per person on average).

**Table 2:**
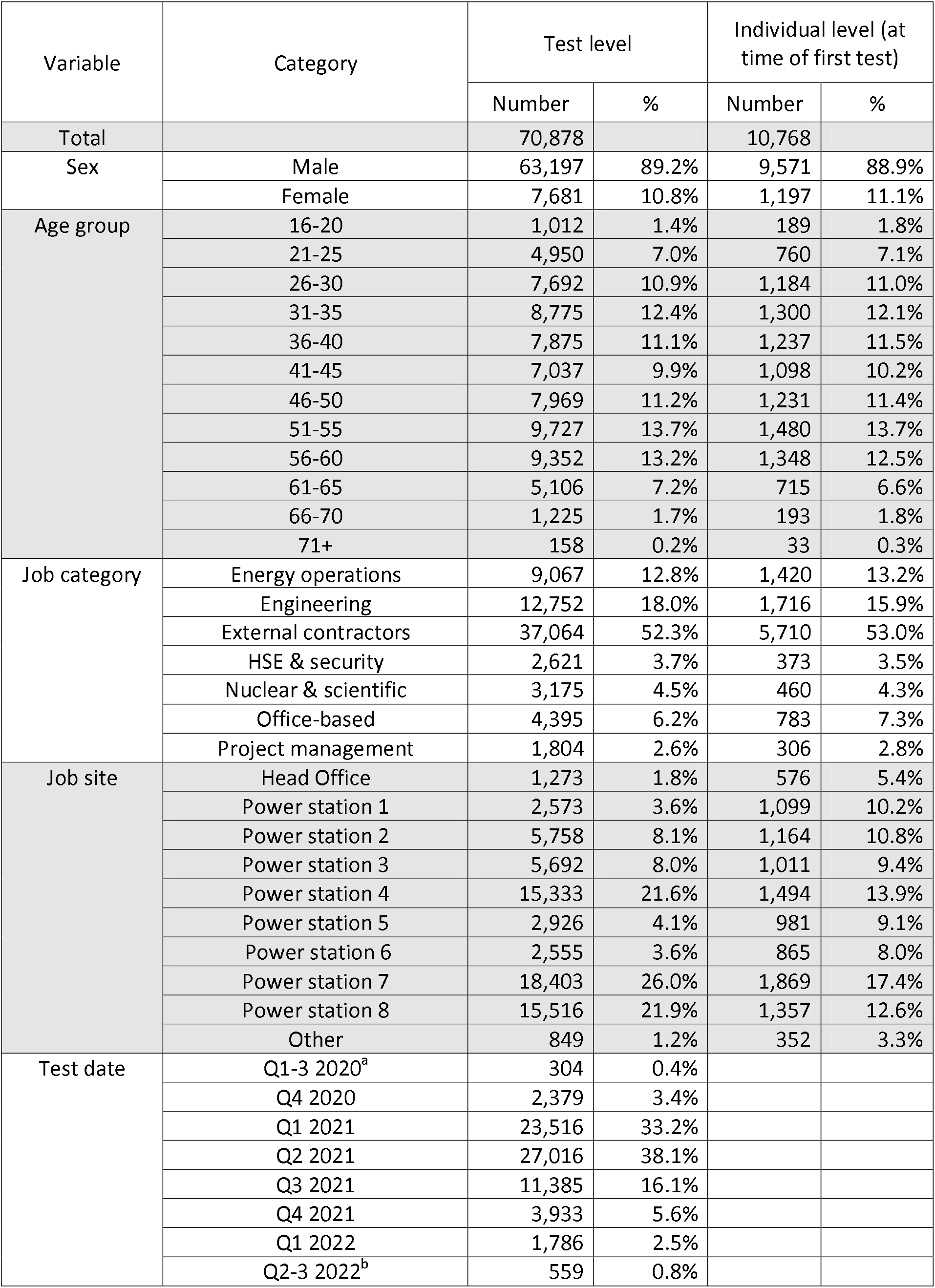
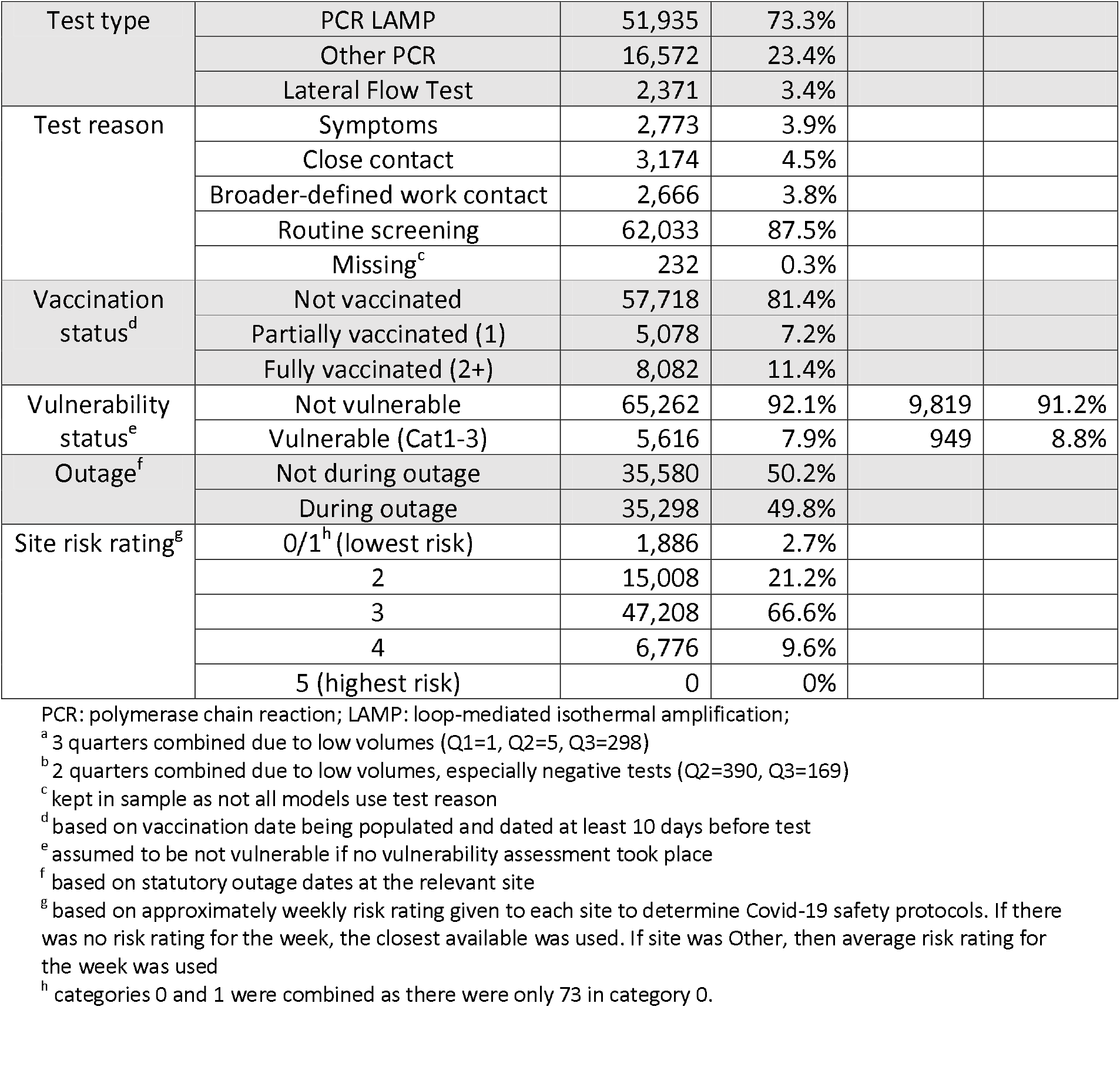
Demographic characteristics of analysis sample.

The number of tests varied hugely by date, reflecting different stages of the pandemic, as well as changes in regulations and protocols of testing and the general prevalence of COVID-19 in the UK. Most tests were in the first half of 2021, but most positive tests were in the first half of 2022 (Figure 1).

**Figure 1:**
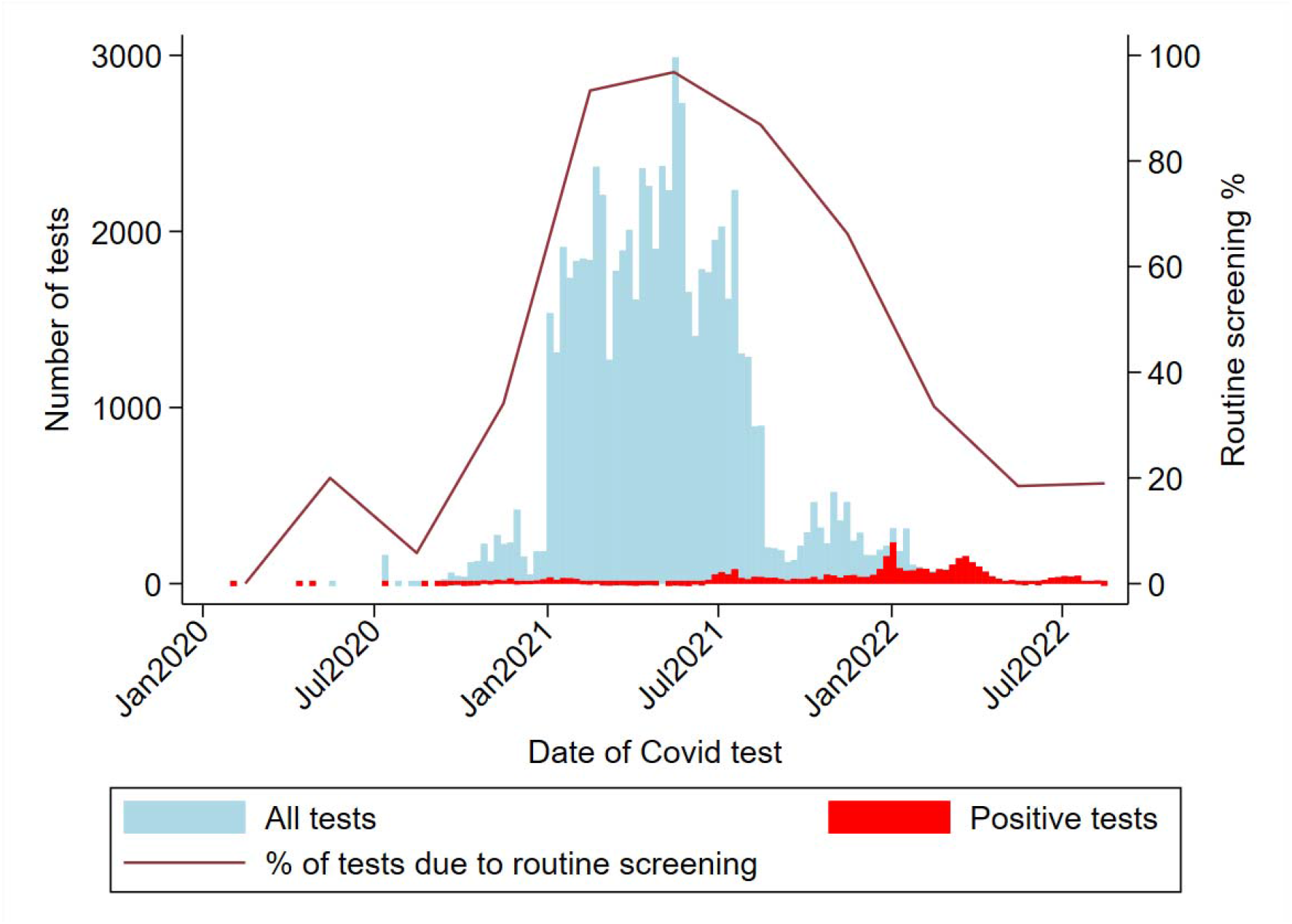
Number of tests, positive tests, and percentage due to routine screening, by date.

The proportions of positive tests that were identified, according to the reasons for testing, are shown in Table 3. Overall, testing those with symptoms identified more than half (54%) of the positive results, whereas testing close contacts picked up 24% and broader-defined work contacts 3%. Nevertheless, a significant minority of cases (16%) were identified by routine screening. The remaining 3% had missing test reason.

**Table 3.**
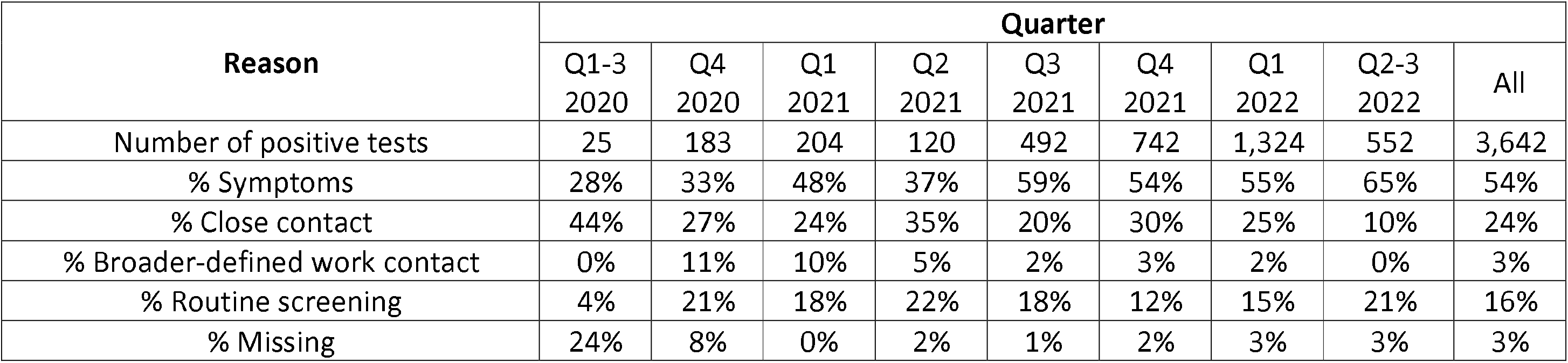
Proportion of positive tests by test reason in each time-period.

| Reason | Quarter |  |  |  |  |  |  |  |  |
| --- | --- | --- | --- | --- | --- | --- | --- | --- | --- |
|  | Q1-3<br>2020 | Q4<br>2020 | Q1<br>2021 | Q2<br>2021 | Q3<br>2021 | Q4<br>2021 | Q1<br>2022 | Q2-3<br>2022 | All |
| Number of positive tests | 25 | 183 | 204 | 120 | 492 | 742 | 1,324 | 552 | 3,642 |
| % Symptoms | 28% | 33% | 48% | 37% | 59% | 54% | 55% | 65% | 54% |
| % Close contact | 44% | 27% | 24% | 35% | 20% | 30% | 25% | 10% | 24% |
| % Broader-defined work contact | 0% | 11% | 10% | 5% | 2% | 3% | 2% | 0% | 3% |
| % Routine screening | 4% | 21% | 18% | 22% | 18% | 12% | 15% | 21% | 16% |
| % Missing | 24% | 8% | 0% | 2% | 1% | 2% | 3% | 3% | 3% |

Table 4 shows the findings for the main risk factors under study. Our “base model” adjusted for test date, age-group and test type (results not shown), as well as the other risk factors shown in the table. There was strong evidence against a linear trend for time-period (p<0.0001) and weak evidence for age group (p=0.05) so these were both included as categorical.

**Table 4:**
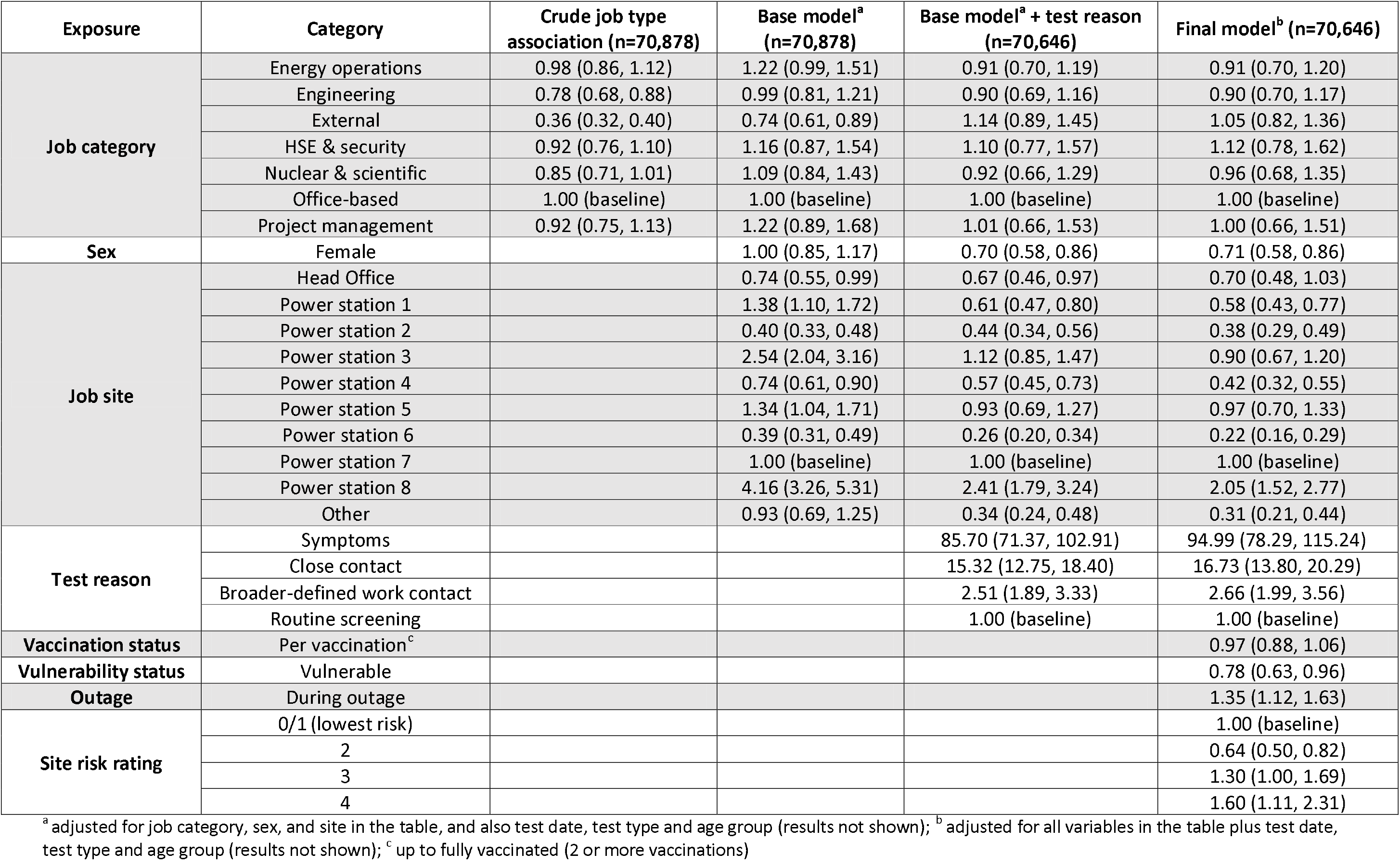
Odds ratios and 95% confidence intervals of risk of testing positive on covid test.

| Exposure | Category | Crude job type association (n=70,878) | Base model<sup>a</sup> (n=70,878) | Base model<sup>a</sup> + test reason (n=70,646) | Final model<sup>b</sup> (n=70,646) |
| --- | --- | --- | --- | --- | --- |
| **Job category** | Energy operations | 0.98 (0.86, 1.12) | 1.22 (0.99, 1.51) | 0.91 (0.70, 1.19) | 0.91 (0.70, 1.20) |
|  | Engineering | 0.78 (0.68, 0.88) | 0.99 (0.81, 1.21) | 0.90 (0.69, 1.16) | 0.90 (0.70, 1.17) |
|  | External | 0.36 (0.32, 0.40) | 0.74 (0.61, 0.89) | 1.14 (0.89, 1.45) | 1.05 (0.82, 1.36) |
|  | HSE & security | 0.92 (0.76, 1.10) | 1.16 (0.87, 1.54) | 1.10 (0.77, 1.57) | 1.12 (0.78, 1.62) |
|  | Nuclear & scientific | 0.85 (0.71, 1.01) | 1.09 (0.84, 1.43) | 0.92 (0.66, 1.29) | 0.96 (0.68, 1.35) |
|  | Office-based | 1.00 (baseline) | 1.00 (baseline) | 1.00 (baseline) | 1.00 (baseline) |
|  | Project management | 0.92 (0.75, 1.13) | 1.22 (0.89, 1.68) | 1.01 (0.66, 1.53) | 1.00 (0.66, 1.51) |
| **Sex** | Female |  | 1.00 (0.85, 1.17) | 0.70 (0.58, 0.86) | 0.71 (0.58, 0.86) |
| **Job site** | Head Office |  | 0.74 (0.55, 0.99) | 0.67 (0.46, 0.97) | 0.70 (0.48, 1.03) |
|  | Power station 1 |  | 1.38 (1.10, 1.72) | 0.61 (0.47, 0.80) | 0.58 (0.43, 0.77) |
|  | Power station 2 |  | 0.40 (0.33, 0.48) | 0.44 (0.34, 0.56) | 0.38 (0.29, 0.49) |
|  | Power station 3 |  | 2.54 (2.04, 3.16) | 1.12 (0.85, 1.47) | 0.90 (0.67, 1.20) |
|  | Power station 4 |  | 0.74 (0.61, 0.90) | 0.57 (0.45, 0.73) | 0.42 (0.32, 0.55) |
|  | Power station 5 |  | 1.34 (1.04, 1.71) | 0.93 (0.69, 1.27) | 0.97 (0.70, 1.33) |
|  | Power station 6 |  | 0.39 (0.31, 0.49) | 0.26 (0.20, 0.34) | 0.22 (0.16, 0.29) |
|  | Power station 7 |  | 1.00 (baseline) | 1.00 (baseline) | 1.00 (baseline) |
|  | Power station 8 |  | 4.16 (3.26, 5.31) | 2.41 (1.79, 3.24) | 2.05 (1.52, 2.77) |
|  | Other |  | 0.93 (0.69, 1.25) | 0.34 (0.24, 0.48) | 0.31 (0.21, 0.44) |
| **Test reason** | Symptoms |  |  | 85.70 (71.37, 102.91) | 94.99 (78.29, 115.24) |
|  | Close contact |  |  | 15.32 (12.75, 18.40) | 16.73 (13.80, 20.29) |
|  | Broader-defined work contact |  |  | 2.51 (1.89, 3.33) | 2.66 (1.99, 3.56) |
|  | Routine screening |  |  | 1.00 (baseline) | 1.00 (baseline) |
| **Vaccination status** | Per vaccination<sup>c</sup> |  |  |  | 0.97 (0.88, 1.06) |
| **Vulnerability status** | Vulnerable |  |  |  | 0.78 (0.63, 0.96) |
| **Outage** | During outage |  |  |  | 1.35 (1.12, 1.63) |
| **Site risk rating** | 0/1 (lowest risk) |  |  |  | 1.00 (baseline) |
|  | 2 |  |  |  | 0.64 (0.50, 0.82) |
|  | 3 |  |  |  | 1.30 (1.00, 1.69) |
|  | 4 |  |  |  | 1.60 (1.11, 2.31) |

<sup>a</sup> adjusted for job category, sex, and site in the table, and also test date, test type and age group (results not shown); <sup>b</sup> adjusted for all variables in the table plus test date, test type and age group (results not shown); <sup>c</sup> up to fully vaccinated (2 or more vaccinations)

We found that the reason for testing was a strong confounder; in particular, the odds ratio for “external” workers (i.e. contractors) changed from 0.74 (95% CI=0.61-0.89) to 1.14 (0.89-1.45) after adjusting for the test reason. Also, women showed lower risks than men, after adjustment for test reason (0.70; 0.58-0.86) but not before 1.00 (0.85-1.17). As well as being a strong confounder of job type, reason for testing was a very strong factor itself (Table 4).

Vulnerability, outages, vaccination status and site risk rating were not identified as confounders (on introduction to the model, results not shown) but were included in the final model for completeness. There was strong evidence against a linear trend for site risk rating (p<0.0001) so this was included as categorical. There was no evidence against a linear trend for vaccine status (p=0.92) so this was included as continuous.

The final model included job type, age, sex, test date, test type, test reason, job site, vaccination status, vulnerability status, outage, and site risk rating. This model showed that there were few differences between job types and likelihood of testing positive after adjusting for all the other included factors. Women were less likely to test positive than men (0.71; 0.58-0.86).

The relationship between site and test positivity showed that power station 8 was higher risk (2.05; 1.52-2.77) than power station 7, the site with the most tests. Power station 6 was the lowest risk (0.22; 0.16, 0.29 compared to power station 7). All other sites were estimated to be similar or lower risk than power station 7 but with varying magnitudes and levels of evidence (Table 4).

There was a large effect of test reason, with those testing due to symptoms having 94.99 (78.29-115.24) times the odds of those from routine screening, those testing due to a positive close contact having 16.73 (13.80-20.29), and broader-defined work contact 2.66 (1.99-3.56) times the odds of those tested in routine screening (Table 4).

There was no evidence of a difference in risk for vaccinated workers (0.97 per vaccination; 0.88-1.06) but vulnerable workers were at lower risk of testing positive than other workers (0.78; 0.63-0.96) and workers testing during an outage were at increased risk (1.35; 1.12-1.63). The site risk rating did not have a linear relationship with an individual worker’s risk of testing positive; category 2 was lower risk than category 1 (baseline), but categories 3 and 4 were higher risk (Table 4).

We have also presented findings stratified by reason for testing in Supplementary Table S1. In general, the findings are consistent with those of the final model (taking into account the wide confidence intervals for some effect estimates). The exception are the findings for those undergoing routine screening, where for some variables the effect estimates were inconsistent with those in the final model. However, once again, the confidence intervals for these effect estimates were relatively wide, due to data sparsity in some time-periods from the changing testing protocols (Figure 1).

## Discussion

As noted in the introduction, there have been relatively few in-depth investigations of work-place risk factors for Covid-19 infection. To our knowledge, no similar study has been done in the electricity-generating industry, so our analyses were largely exploratory, with the aim of assessing a number of potential risk factors for infection in this context. This industry is of particular interest, since working from home is difficult and unusual, and there is relatively little contact with the general public during working hours. It thus provides an opportunity to study workplace risk factors for transmission in this environment.

In this test-negative design study, based on data from a United Kingdom electricity-generating company, we estimated the odds ratios for infection by job category, site, reason for testing, vulnerability, sex, reason for testing and the COVID-19 weekly risk rating for each site, adjusting for age and test date.

With regards to ‘reason for testing’, it is partly because of this variable that we used the test-negative design. For example, if one particular job primarily involves routine testing, whereas another job primarily involves testing only when symptoms occur, we would expect to find differences between these two jobs in the rate of positive tests. One of the co-authors (NP) has written on precisely this problem in a follow-up[20] to a paper on the use of the TND for analysing Covid-19 test results[22], stating: “For the application of the TND to study risk factors in situations where testing includes symptomatic as well as non-symptomatic persons, the reason for testing is important to record and account for in analysis and inference… increasingly people are being tested for a variety of reasons, and it is therefore necessary to control for ‘reason for testing’ in the analysis.”

There was little difference in risk by job category after adjusting for other factors. There were large differences in risk between job sites which could be for a variety of reasons including localised community infection rates (partially adjusted for by site risk rating), whether the site was always operational (some weren’t), and differences in workplace culture. In general, the electricity generating sites include in this study were high hazard, critical national infrastructure, highly regulated and with a strong safety culture. Women were less likely to be infected than men (OR=0.71). The strongest predictor of a positive test in our data was the reason for testing. For example, someone who was tested because of symptoms had 95 times the odds of testing positive compared to someone who was routinely screened. Those who were tested because of close contact with a case, had 17 times the odds of testing positive, and those with broader-defined work contact had 2.7 times the odds. This is consistent with the Covid-19 literature – unsurprisingly, those with symptoms are much more likely to have an infection than randomly selected people without symptoms. Thus, this is not an original finding, but it does reinforce the importance of adjusting for reason for testing in the analyses. Reason for testing was also a strong confounder for job category, sex and job site. (Table 4)

It is also notable, that despite these strong associations with symptomatic and contact testing, across the pandemic, 16% of cases were identified by routine testing. Thus, routine testing may have played an important role in identifying a large minority of cases, and thereby also reducing the spread of infection to contacts.

One limitation of this study is that we have included multiple tests on the same person in each time-period (Supplementary Table S2). If the tests are well-spaced then they should still be independent. However, tests close together on the same person will be more likely to have the same result as each other. One option is to shorten each time-period and remove multiple tests, but if the time-periods are very short then there are problems of sparse data due to the large number of parameters in the model. In addition, although most individuals had up to 4 tests in a time-period, there were some with substantially more. These could bias results towards the null as there is a maximum of one positive result per person and so the multiple negative tests would outweigh the positive one when estimating the effects.

Overall, these findings showed little difference in positivity rates by job category once the analyses were adjusted for test reason. There were some differences by site, with four sites showing substantially lower risks, and one site showing higher risks in the final model. Vulnerable individuals showed slightly lower risks, possibly due to those individuals taking more care. Positivity rates were slightly higher during outages when there could be a lot more people on site. Vaccination did not show a protective effect on testing positive which is perhaps surprising, and also in contrast with results from Rhodes et al, which showed that number of vaccines was inversely related to infection risk in working age people taking part in the Covid-19 infection survey in the UK.[23] Only about a fifth of tests in the present study related to a worker that was vaccinated at the time of testing, as vaccines for the majority of workers were not introduced until the middle of 2021.

The site risk rating showed a consistent (but modest) dose-response with infection risk, indicating that such risk ratings may be useful for identifying “high risk” sites. It could be argued that the site risk rating depends on the number of cases so it may not be appropriate to always adjust for it. However, inclusion of this variable made little difference to the main effect of job category (Table 4) and the relationship between site risk rating and the odds of testing positive is of interest in itself.

## Data Availability

Data may be obtained from a third party and are not publicly available.

## Research Ethics Approval

The Observational Research Committee of the London School of Hygiene and Tropical Medicine gave ethical approval for this work (ref 28125).

## Copyright

© Crown copyright (2024) Health and Safety Executive. This is an open access article distributed under the terms of the Open Government Licence v3.0, which permits re-use, distribution, reproduction, and adaptation, provided the original work is properly cited.

