## Supplementary Table for "Risk factors for SARS-Cov-2 infection at a United Kingdom electricity-generating company: a test-negative design case-control study"

### Supplementary Tables

**Table S1: Fully adjusted odds ratios and 95% confidence intervals from separate logistic regression models for each test reason**

| Exposure | Category | Final model[a] (n=70,686) | Symptoms[b] (n=2,772[c]) | Close contacts[b] (n=3175) | Broader-defined work contacts[b] (n=2,599[d]) | Routine screening[b] (n=61,764[e]) |
|---|---|---|---|---|---|---|
| Job category | Energy operations | 0.91 (0.70, 1.20) | 0.94 (0.59, 1.50) | 0.69 (0.42, 1.13) | 0.34 (0.14, 0.87) | 1.87 (1.06, 3.31) |
| | Engineering | 0.90 (0.70, 1.17) | 1.08 (0.69, 1.70) | 0.79 (0.48, 1.28) | 0.27 (0.10, 0.73) | 1.12 (0.65, 1.93) |
| | External | 1.05 (0.82, 1.36) | 1.20 (0.77, 1.88) | 1.55 (0.96, 2.51) | 0.36 (0.14, 0.93) | 0.93 (0.55, 1.59) |
| | HSE & security | 1.12 (0.78, 1.62) | 0.77 (0.40, 1.49) | 1.07 (0.56, 2.05) | 0.79 (0.28, 2.20) | 1.54 (0.73, 3.24) |
| | Nuclear & scientific | 0.96 (0.68, 1.35) | 0.76 (0.43, 1.34) | 1.09 (0.56, 2.12) | 0.10 (0.01, 0.83) | 1.78 (0.92, 3.45) |
| | Office-based | 1.00 (baseline) | 1.00 (baseline) | 1.00 (baseline) | 1.00 (baseline) | 1.00 (baseline) |
| | Project management | 1.00 (0.66, 1.51) | 0.67 (0.33, 1.35) | 1.16 (0.51, 2.64) | 0.62 (0.17, 2.26) | 1.75 (0.80, 3.83) |
| Sex | Female | 0.71 (0.58, 0.86) | 0.50 (0.35, 0.72) | 0.83 (0.57, 1.22) | 0.86 (0.40, 1.87) | 0.96 (0.65, 1.41) |
| Job site | Head Office | 0.70 (0.48, 1.03) | 0.53 (0.23, 1.20) | 0.58 (0.25, 1.36) | 0.13 (0.00, 3.69) | 0.60 (0.32, 1.14) |
| | Power station 1 | 0.58 (0.43, 0.77) | 0.27 (0.13, 0.56) | 0.25 (0.15, 0.43) | 0.18 (0.03, 1.25) | 2.05 (1.17, 3.59) |
| | Power station 2 | 0.38 (0.29, 0.49) | 0.18 (0.10, 0.34) | 0.19 (0.11, 0.33) | 0.11 (0.02, 0.65) | 1.15 (0.71, 1.86) |
| | Power station 3 | 0.90 (0.67, 1.20) | 0.24 (0.12, 0.46) | 0.86 (0.50, 1.48) | 0.11 (0.02, 0.71) | 1.48 (0.85, 2.58) |
| | Power station 4 | 0.42 (0.32, 0.55) | 0.13 (0.07, 0.25) | 0.19 (0.11, 0.33) | 0.13 (0.02, 0.78) | 1.39 (0.86, 2.24) |
| | Power station 5 | 0.97 (0.70, 1.33) | 0.41 (0.19, 0.89) | 0.51 (0.27, 0.97) | 0.10 (0.01, 0.78) | 2.93 (1.72, 4.98) |
| | Power station 6 | 0.22 (0.16, 0.29) | 0.12 (0.06, 0.24) | 0.20 (0.12, 0.34) | 0.04 (0.01, 0.28) | 0.21 (0.12, 0.38) |
| | Power station 7 | 1.00 (baseline) | 1.00 (baseline) | 1.00 (baseline) | 1.00 (baseline) | 1.00 (baseline) |
| | Power station 8 | 2.05 (1.52, 2.77) | 1.17 (0.50, 2.75) | 3.90 (1.96, 7.74) | 0.84 (0.03, 27.46) | 2.09 (1.24, 3.53) |
| | Other | 0.31 (0.21, 0.44) | 0.13 (0.06, 0.28) | 0.18 (0.10, 0.34) | 0.00 (0.00, 0.05) | 0.94 (0.44, 2.01) |
| Vaccination status | Per vaccination[f] | 0.97 (0.88, 1.06) | 0.89 (0.76, 1.06) | 0.86 (0.72, 1.02) | 0.80 (0.52, 1.22) | 0.89 (0.75, 1.05) |
| Vulnerability status | Vulnerable | 0.78 (0.63, 0.96) | 0.87 (0.56, 1.35) | 1.43 (0.99, 2.05) | 0.57 (0.24, 1.34) | 0.87 (0.56, 1.35) |
| Outage | During outage | 1.35 (1.12, 1.63) | 0.54 (0.39, 0.74) | 5.15 (3.44, 7.71) | 4.10 (1.41, 11.94) | 0.54 (0.39, 0.74) |
| Site risk rating | 0/1 (lowest risk) | 1.00 (baseline) | 1.00 (baseline) | 1.00 (baseline) | 1.00 (baseline) | 1.00 (baseline) |
| | 2 | 0.64 (0.50, 0.82) | 0.37 (0.23, 0.62) | 1.15 (0.76, 1.74) | 1.36 (0.40, 4.65) | 0.37 (0.23, 0.62) |
| | 3 | 1.30 (1.00, 1.69) | 1.05 (0.62, 1.78) | 2.10 (1.36, 3.23) | 4.05 (1.26, 13.02) | 1.05 (0.62, 1.78) |
| | 4 | 1.60 (1.11, 2.31) | 0.51 (0.24, 1.10) | 2.74 (1.44, 5.22) | 5.18 (1.11, 24.15) | 0.51 (0.24, 1.08) |

[a] adjusted for all variables in the table plus test date, test type, age and test reason; [b] adjusted for all variables in the table plus test date, test type, and age;
[c] one observation omitted; [d] 64 observations omitted; [e] 269 observations omitted; [f] up to fully vaccinated (2 or more vaccinations);

**Table S2: Frequency of number of tests per person in each time-period in the model**

| Number of tests per person | Q1-3 2020 | Q4 2020 | Q1 2021 | Q2 2021 | Q3 2021 | Q4 2021 | Q1 2022 | Q2-3 2022 | Overall |
|---|---|---|---|---|---|---|---|---|---|
| 1 | 220 (84%) | 911 (59%) | 2,228 (37%) | 1,504 (24%) | 1,641 (41%) | 1,401 (63%) | 1,500 (92%) | 557 (100%) | 2,551 (24%) |
| 2 | 39 (15%) | 492 (32%) | 940 (15%) | 833 (13%) | 671 (17%) | 420 (19%) | 129 (8%) | 1 (0%) | 1,403 (13%) |
| 3 | 2 (1%) | 79 (5%) | 498 (8%) | 591 (9%) | 409 (10%) | 169 (8%) | 8 (0%) | | 932 (9%) |
| 4 | | 39 (3%) | 321 (5%) | 502 (8%) | 348 (9%) | 95 (4%) | 1 (0%) | | 707 (7%) |
| 5 | | 7 (0%) | 298 (5%) | 546 (9%) | 307 (8%) | 51 (2%) | | | 528 (5%) |
| 6 | | 4 (0%) | 371 (6%) | 667 (11%) | 284 (7%) | 36 (2%) | | | 473 (4%) |
| 7 | | 2 (0%) | 370 (6%) | 826 (13%) | 191 (5%) | 23 (1%) | | | 408 (4%) |
| 8 | | 1 (0%) | 294 (5%) | 497 (8%) | 88 (2%) | 12 (1%) | | | 349 (3%) |
| 9 | | | 232 (4%) | 142 (2%) | 31 (1%) | 4 (0%) | | | 345 (3%) |
| 10 | | 1 (0%) | 202 (3%) | 59 (1%) | 10 (0%) | 3 (0%) | | | 336 (3%) |
| 11 | | | 211 (3%) | 63 (1%) | 4 (0%) | 1 (0%) | | | 311 (3%) |
| 12 | | | 86 (1%) | 42 (1%) | 2 (0%) | | | | 312 (3%) |
| 13 | | | 26 (0%) | 19 (0%) | 2 (0%) | | | | 314 (3%) |
| 14 | | | 7 (0%) | 12 (0%) | 1 (0%) | | | | 314 (3%) |
| 15 | | | 5 (0%) | 4 (0%) | | | | | 317 (3%) |
| 16 | | | | 1 (0%) | 1 (0%) | | | | 292 (3%) |
| 17 | | | | | | | | | 233 (2%) |
| 18 | | | | | | | | | 192 (2%) |
| 19 | | | | 1 (0%) | | | | | 134 (1%) |
| 20+ | | | | | | | | | 317 (3%) |
